# Rapid “mix and read” assay for scalable detection of SARS-CoV-2 antibodies in patient plasma

**DOI:** 10.1101/2020.09.01.20184101

**Authors:** Hong Yue, Radosław P. Nowak, Daan Overwijn, N. Connor Payne, Stephanie Fischinger, Caroline Atyeo, Lindsey R. Baden, Eric J. Nilles, Elizabeth W. Karlson, Xu G. Yu, Jonathan Z. Li, Galit Alter, Ralph Mazitschek, Eric S. Fischer

## Abstract

The human beta coronavirus SARS-CoV-2, causative virus of COVID-19, has infected more than 15 million people globally and continues to spread. Widespread, population level testing to detect active and past infections is critical to curb the COVID-19 pandemic. Antibody (serological) testing is the only option for detecting past infections outside the narrow window accessible to nucleic acid-based tests. However, currently available serological assays commonly lack scalability. Here, we describe the development of a rapid homogenous serological assay for the detection of antibodies to SARS-CoV-2 in patient plasma. We show that the fluorescence-based assay accurately detects seroconversion in COVID-19 patients from less than 1 *μ*L of plasma. Using a cohort of samples from COVID-19 infected or healthy individuals, we demonstrate detection with 100% sensitivity and specificity. This assay addresses an important need for a robust, low barrier to implementation, and scalable serological assay with complementary strengths to currently available serological platforms.

## Introduction

SARS-CoV-2 (severe acute respiratory syndrome coronavirus-2) is a human beta-coronavirus that has caused a global pandemic of coronavirus disease 2019 (COVID-19)^1^. To better understand the spread of the disease and manage public health responses, rapid and ideally population wide screening is essential. Nucleic acid-based tests for the identification of infected individuals have been widely implemented^2^, but these tests can only detect the virus during a narrow window of acute disease. Robust serological assays are necessary to identify individuals who have previously been infected and developed antibodies to SARS-CoV-2. Such serological assays for SARS-CoV-2 antibodies are also critical to identify individuals with asymptomatic disease^3, 4^. Serological testing can establish the spread of the virus within a population and help epidemiologists to accurately model the prevalence of COVID-19. Important to the large number of efforts to develop SARS-CoV-2 vaccines, robust serological testing assays provide means to monitor level and duration of response in clinical trials as well as trace serological response in the general population post-vaccination.

The serological assays currently used to detect anti-SARS-CoV-2 antibodies are either enzyme-linked immunosorbent assays (ELISA) or commercial solutions on large diagnostics platforms^3, 5-12^. However, these strategies have several critical limitations^13-17^. ELISA assays suffer from limited scalability, mainly due to multi-step protocols with extensive wash steps that lead to the need for specialized equipment and automation. Other available assays are not quantitative and require specialized analytical laboratory platforms that are not widely available. To address these limitations, we developed a robust, “mix and read” assay for the detection of SARS-CoV-2 antibodies based on time-resolved Förster resonance energy transfer (TR-FRET). Given its homogenous nature, the assay is highly scalable and does not require complex automation. In direct comparison with the current state-of-the-art ELISA or commercial assays, our TR-FRET assay platform performs with comparable or superior specificity and sensitivity, while the processing time is reduced to ~ 1 hour compared to several hours for ELISA. Importantly, the lack of multiple steps, including extensive washes, results in superior repeatability and reproducibility and makes the assay suitable for manual operation as well as highly scalable automation.

## Results

### Development of a TR-FRET assay to detect SARS-CoV-2 antibodies

We report the development and validation of a homogenous serological assay platform for the detection of SARS-CoV-2 antibodies in human plasma/serum (**Fig. 1a**). The direct detection of a ternary complex between antigen and serum antibodies using a TR-FRET readout allows for a simple mix and read protocol that lends itself to scalable automation (**Fig. 1b**). The proximity of the donor (Terbium) and acceptor (BODIPY) fluorophores induced by presence of serum antibodies results in a positive TR-FRET signal that is read out as a 520 nm (acceptor) / 490 nm (donor) ratio and allows for accurate quantification of serum antibodies in an isotype specific manner (**Fig. 1a**).

**Fig. 1.**
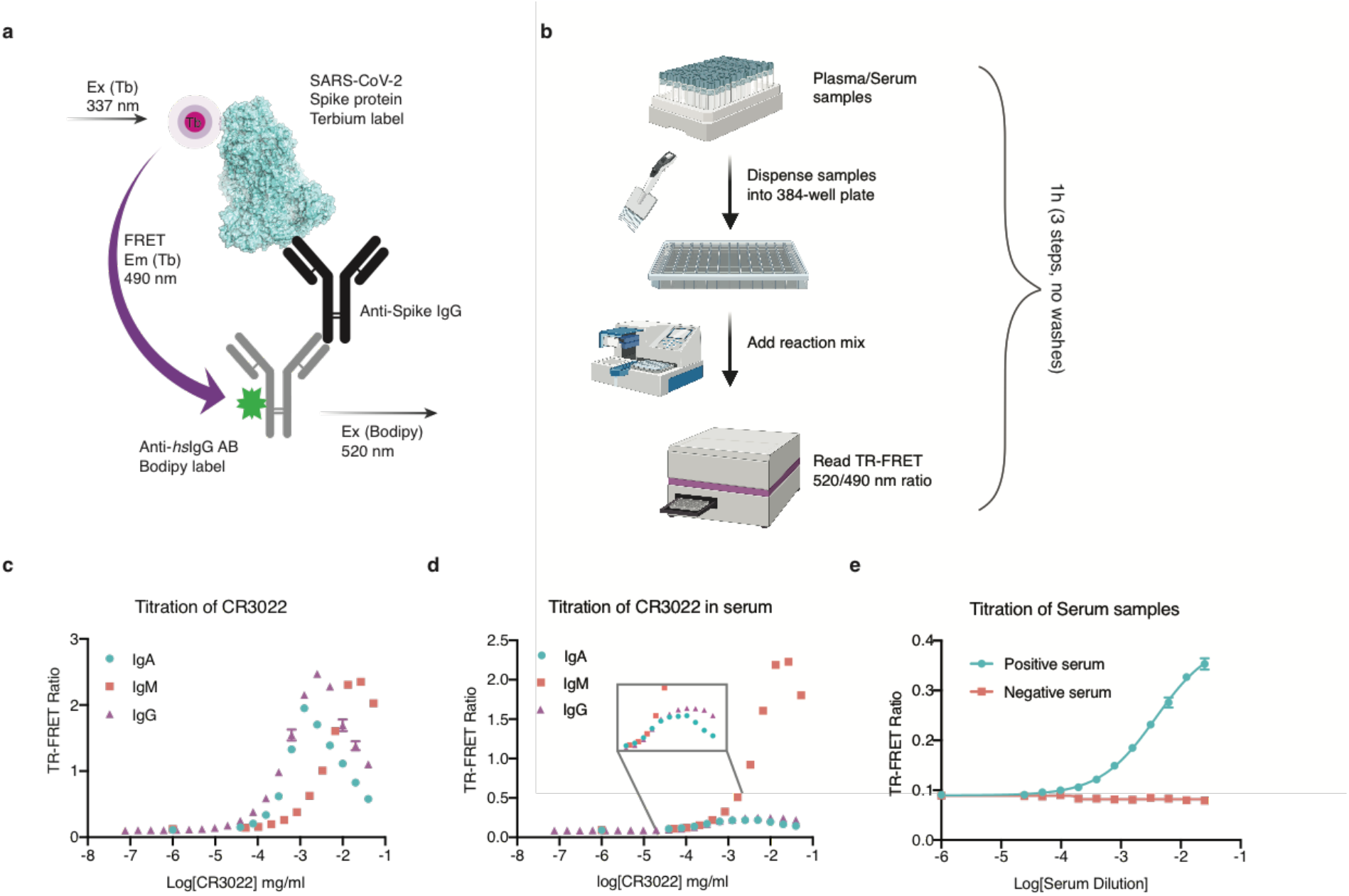
Assay setup and CR3022 validation. **a**, Principle of TR-FRET assay. Antibodies recognizing human IgG are labeled with BODIPY. SARS-CoV-2 S protein is labeled with Tb (Terbium) and both are mixed with serum for isotype specific antibody detection. The light pulse at 337 nm excites Terbium chelate of Tb-S labeled protein and emits light at 490 nm which in turn excites BODIPY-labeled secondary antibodies found in proximity induced by analyte generating a TR-FRET signal detected at 520 nm. **b**, Flow-chart of TR-FRET assay. The serum samples are diluted into multiple well plates and added into reaction mixture. Reaction mixture is added beforehand by automated dispenser (Multidrop Combi Reagent Dispenser). The diluted serum samples are added into the reaction mixture using a Crystal Gryphon (Art Robbins Instruments, LLC) or manually by multichannel pipetting. Plates are read on TR-FRET compatible plate reader (PHERAstar FSX Microplate Reader). **c**, Titration of CR3022 IgG/IgM/IgA1 into preformed mix of Tb-S protein (7.5 nM final) and BODIPY labelled αIgG/αIgM/αIgA (250 nM final). **d**, as in panel c, but in presence of 1:150 dilution of negative serum. Data of Fig. 1c and Fig. 1d are represented as means ± SD of two replicates (n = 2). **e**, titration of positive and negative serum in final assay condition 250 nM BODIPY-αIgG and 7.5 nM Tb-S. Data in Fig. S2 are represented as means ± SD of three replicates (n = 3).

To enable sensitive detection, we optimized assay conditions to minimize the signal-to-noise ratio as follows. We first established that the TR-FRET assay format can detect the binding of immunoglobulin variants IgG, IgM and IgA1 to SARS-CoV-2 antigens (**Fig. 1c**). The SARS-CoV-2 spike protein (S protein) is responsible for binding to the host receptor ACE2 to mediate virus entry upon infection^18^ and most neutralizing antibodies have been found to target the S protein^19^. We therefore used S protein and the receptor binding domain of the SARS-CoV-2 spike protein (S-RBD) for assay development. S protein and S-RBD, expressed and purified from Chinese hamster ovarian cells (CHO), were labeled with Terbium or BODIPY (see **methods**). Detection antibodies (αIgG, αIgM, αIgA1) were commercially obtained and also labeled with either Terbium or BODIPY. As positive control, we used recombinantly expressed SARS-1 IgG antibody CR3022^20^ that has been shown to cross-react with the RBD of SARS-CoV-2 (K_d_ of 9.1 ± 0.7 nM, **Fig. S1a**) and IgM and IgA1 antibodies engineered to contain the CR3022 variable region^21^. To identify ideal labeling positions, titrations of CR3022 (IgG, IgM, IgA1) into a mix of labeled S-RBD and labeled detection antibody were performed while varying the position of donor and acceptor (either on S-RBD or detection antibody) (**Fig. S1b, c)**. While we found that all combinations lead to a functional readout (**Fig. S1d, e**), we observed that terbium conjugation to the antigen results in optimal performance when used with serum/plasma samples. We therefore selected labeling the antigen with terbium and the detection antibody with BODIPY, resulting in quantitative binding curves for CR3022 (IgG/IgM/IgA1) (**Fig. 1c**). The binding curves exhibit the characteristic bell-shape due to the prozone effect^22^, which can be accurately accounted for by mathematical models^23^. Next, we established that CR3022 can similarly be detected in human serum (**Fig. 1d**), and while the signal is reduced, the low background level allows for accurate quantification. During the optimization of assay conditions, we noticed that replacing RBD with the full-length SARS-CoV-2 spike protein (S protein) significantly reduced background particularly in the presence of serum. This is likely due to 1) the trimeric nature of the full-length S protein leading to avidity effects; and 2) the exposure of otherwise shielded RBD surfaces that may cause unspecific interactions. We therefore continued all further validation using S protein and subsequently optimized the concentrations of antigen and detection antibody (**Fig. S2a, b**). With optimized concentrations, we validated that convalescent serum results in a dose dependent response in TR-FRET signal (**Fig. 1e**).

### Homogenous TR-FRET assay can detect IgG in patient serum

To test the detection of antibodies in serum obtained from positive and negative controls, a set of 49 PCR tested positive (CoV2+) patients from Mass General and Brigham and Women’s Hospital, 28 PCR tested negative (healthy, CoV2-) patients from Mass General Hospital, as well as 19 pre-pandemic serum samples from Mass General Brigham Biobank (CoV2-) or community volunteers (CoV2-) was assembled (hereafter referred to as 96w_testset). Adapting established protocols^24^, we performed an ELISA assay using S protein as reference (**Fig. 2a**). We next profiled the 96w_testset with our TR-FRET assay at an initial serum dilution of 1:100 to match the exact ELISA concentration (**Fig. 2b**), and found that using a 3 standard deviations (SD) cutoff away from the healthy control mean, the TR-FRET achieves 94.9% sensitivity and 100% specificity while the ELISA achieves 100% sensitivity and 96.6% specificity. We observe a strong correlation between the TR-FRET and ELISA assays (**Fig. 2c**). While the discrimination between CoV2+ and CoV2-was comparable between TR-FRET and ELISA, we found that the ELISA had stronger signal compared to TR-FRET especially for low responders. This can be explained by the signal amplification in the ELISA compared to equilibrium binding of the TRFRET assay. However, the lower signal is offset by the low background noise of the TR-FRET. We nevertheless wondered whether signal could further be boosted without compromising background noise by increasing the serum concentration, and how the assay would react to changes in serum dilution. We re-assayed the 96w_testset using dilution factors of 1:150 or 1:50 and observed that increasing the serum concentration improves signal strength without compromising background noise (**Fig. 2d, e**). Lowering the serum concentration to 1:150 only marginally reduces performance (**Fig. 2f, g**). When analyzing this initial 96w_testset, we observed several CoV2+ samples with unexpected low response. We hypothesized that this might be the result of epitope masking since the TR-FRET assay utilizes covalent labeling of the antigen with terbium. We therefore sought to ensure minimal epitope masking and optimized the Degree of Labelling (DOL) (**Fig. S3)**. We found that a DOL ~ 3.8 resulted in no detectable epitope masking with optimal signal and therefore a DOL ~ 3.8 was ensured for all further experiments.

**Fig. 2.**
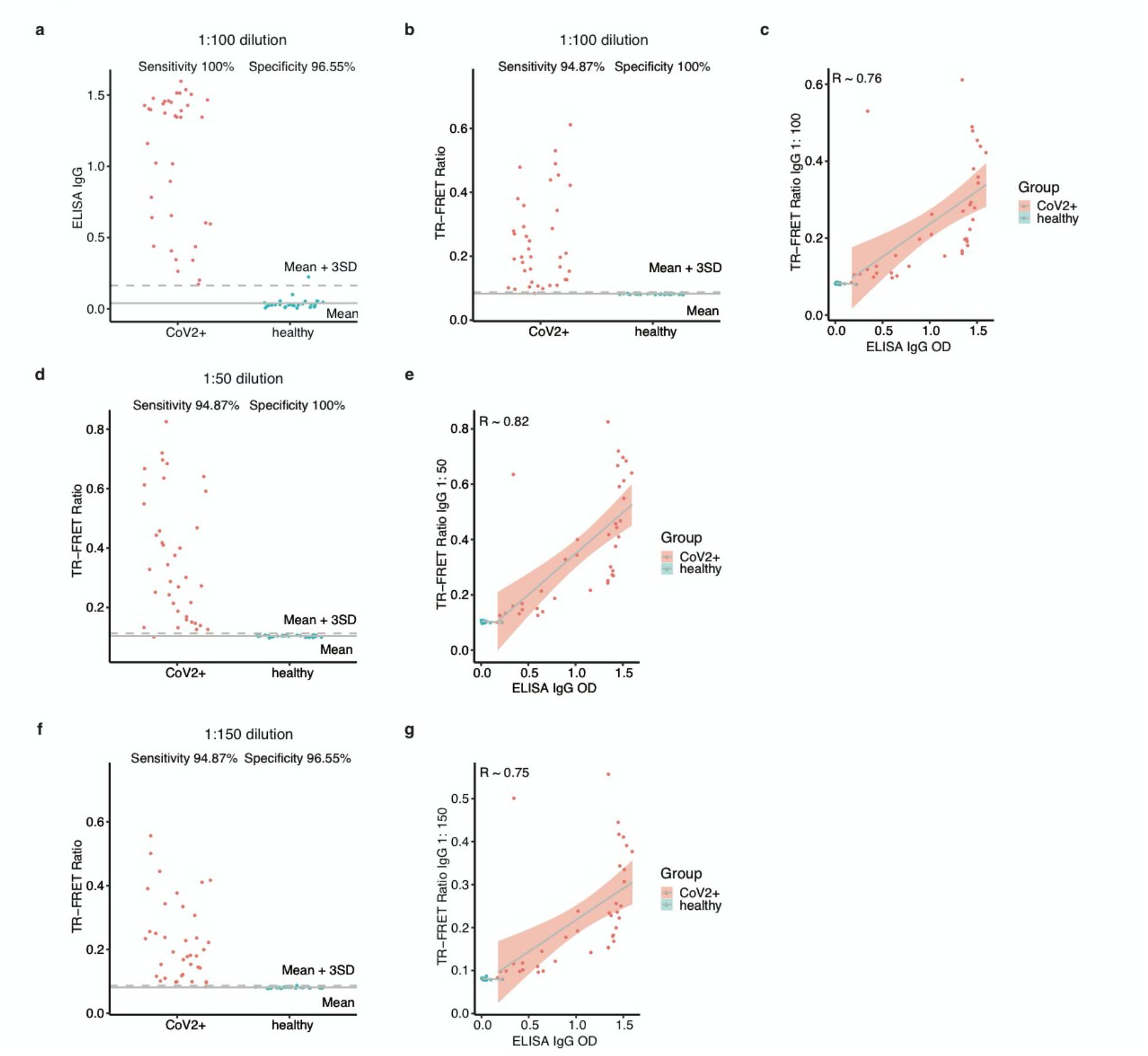
Optimization of a serum dilution factor. **a**, Sensitivity and specificity of ELISA IgG assay. Serum samples from a cohort of 49 PCR positive and 28 PCR negative samples (96w_testset) were diluted at **1:100** serum to buffer ratios and processed in ELISA assay, **b**, as in panel a, but for TR-FRET αIgG-S assay on the same cohort at **1:100** dilution, **c**, Correlation of TR-FRET αIgG-S assay with serum dilution 1:100 and the ELISA αIgG-S at 1:100 serum dilution. **d**, Sensitivity and specificity analysis for TR-FRET αIgG-S assay with the 1:50 serum dilution. **e**, Correlation of TR-FRET αIgG-S assay with serum dilution 1:50 and the ELISA αIgG-S at 1:100 serum dilution. **f**, Sensitivity and specificity analysis for TR-FRET αIgG-S assay with the 1:150 serum dilution. **g**, Correlation of TR-FRET αIgG-S assay with serum dilution 1:150 and the ELISA αIgG-S at 1:100 serum dilution. Data are represented as means ± SD of three replicates (n = 3).

### TR-FRET assay can accurately detect seroconversion

With optimized conditions and DOL, we next used the TR-FRET assay to detect seroconversion in a larger set of samples from the Mass General Brigham Biobank containing 68 SARS-CoV-2 PCR positive samples (CoV2+), and 100 pre-pandemic healthy controls (CoV2-) (hereafter referred to as MGB set) that was again profiled using the established ELISA assay for reference and has also been profiled with different commercial and academic assays^25^. In line with our previous observations, the standard deviation of the healthy controls was very low, and accurate discrimination between CoV2+ and healthy samples was achieved with 100% specificity and 100% sensitivity using a cut-off based on 3 SD of the healthy control (**Fig. 3a**). The results are comparable to the established ELISA assay on the same sample set (**Fig. 3b**), and the response of individual samples between TR-FRET and ELISA assays was well correlated (Pearson correlation coefficient of 0.91) (**Fig. 3c**).

**Fig. 3.**
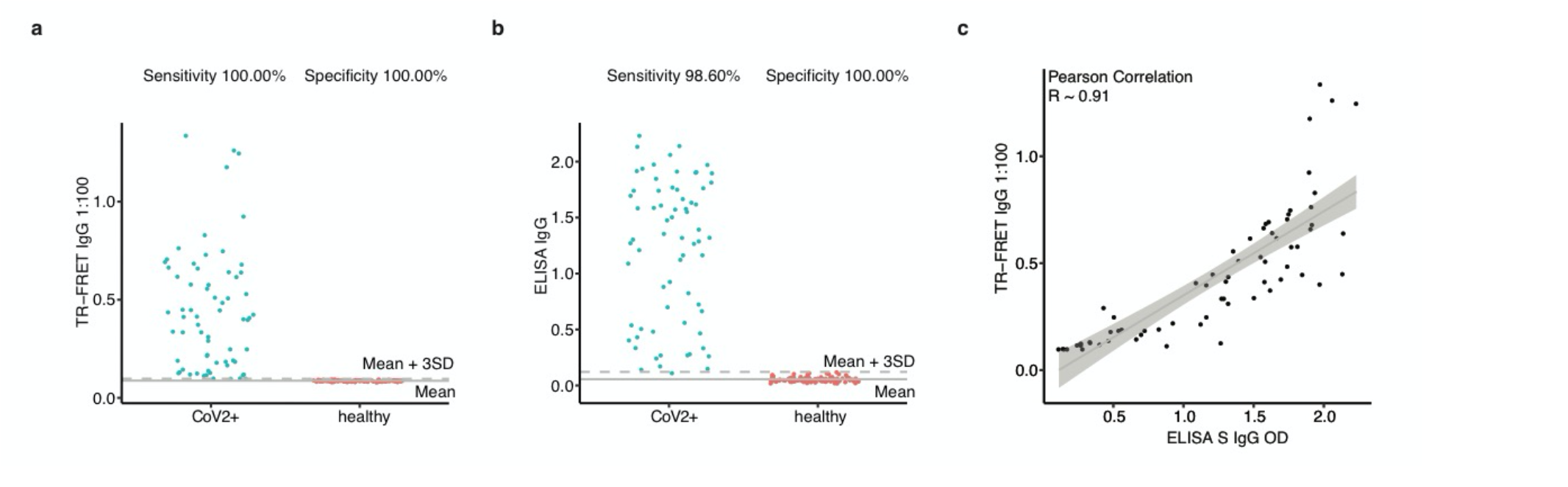
Sensitivity and specificity. **a**, Sensitivity and specificity of TR-FRET αIgG-S assay performed on a cohort of 68 SARS-CoV-2 PCR positive samples (CoV2+), and 100 prepandemic negative samples (healthy) **b**, Sensitivity and specificity of ELISA IgG performed on the same cohort, **c**, Correlation of TR-FRET IgG and ELISA IgG at the same concentration of serum. Data are represented as means ± SD of two replicates (n = 2).

### TR-FRET assay analytical limit of detection

In order to assess the limit of detection (LoD) of the TR-FRET assay, we first performed a titration of control antibody CR3022 IgG in the presence and absence of negative control serum at 1:150 serum to buffer dilution (**Fig. 4a**). The prozone effect is clearly visible at higher concentrations of the antibody and signal intensity is reduced in the presence of serum. We next selected the lowest concentrations of CR3022 IgG antibody where signal was higher than mean + 3 SD and compared 20 replicates against the 20 replicates of blank control both in presence and absence of serum (**Fig, 4b**). Based on this, the LoD for the TR-FRET assay was determined to be 1.22 ng/mL in absence of serum, and 39 ng/mL in presence of the serum, which is in the range of common ELISA LoD ^26^.

**Fig. 4.**
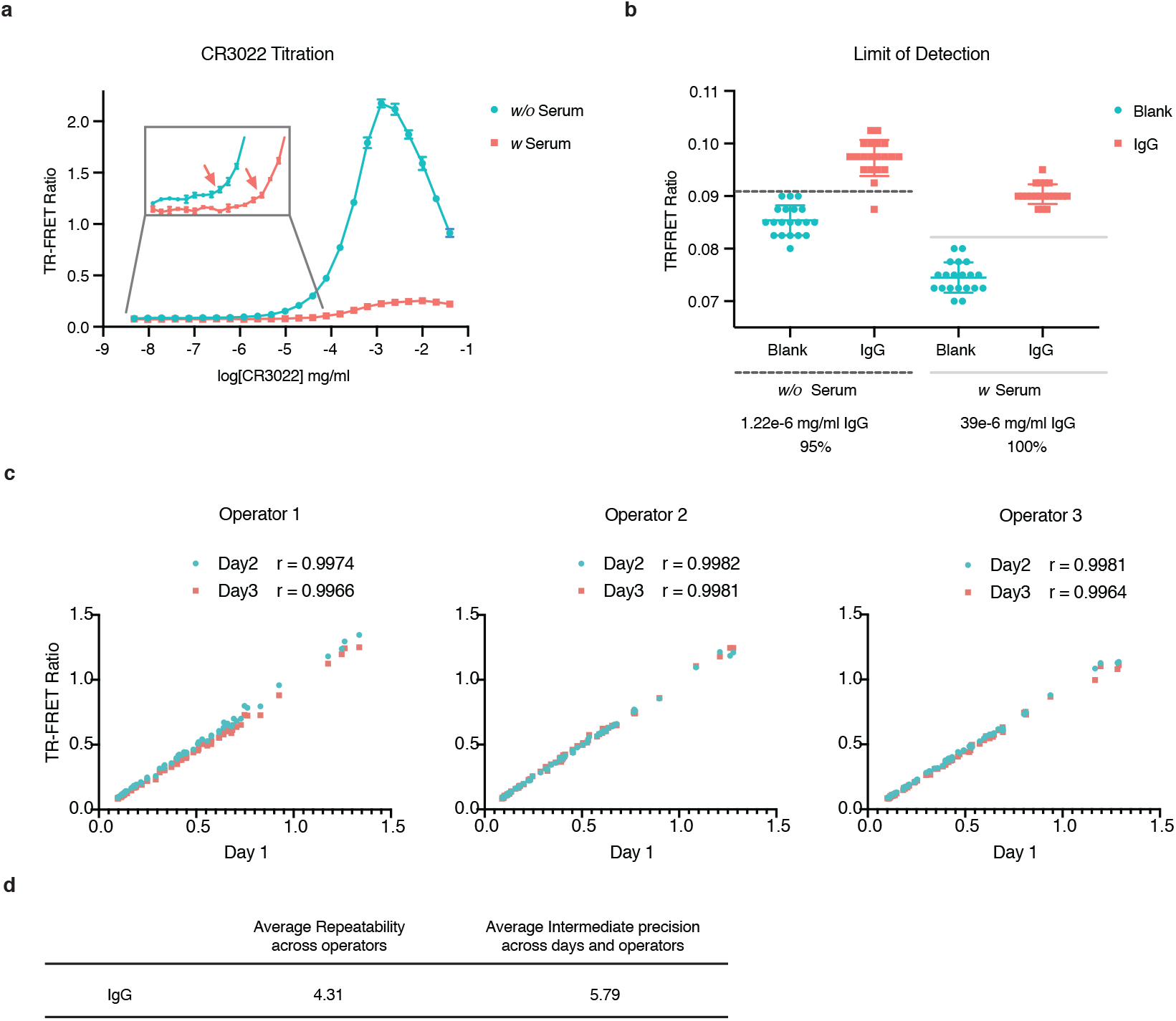
Precision and Limit of Detection. **a**, TR-FRET αIgG-S assay. Titration of CR3022 IgG in presence and absence of negative serum at 1:100 dilution. Data are represented as means ± SD of three replicates (n = 3). The concentration of CR3022 selected for LoD study is highlighted with red arrow. **b**, TR-FRET αIgG-S. Limit of detection for TR-FRET assay was assessed by comparing 20 replicates (n = 20) of the CR3022 to 20 replicates of buffer control (n = 20) in presence and absence of negative serum at 1:100 dilution. **c**, Comparison between three independent runs of performed on different days by three different operators of a TR-FRET αIgG-S assay on a set of positive responders as well as negative control samples (68 total). **d**, the calculated average repeatability across operators (CV%) and average intermediate precision (calculated across days and operators) corresponding to data in Fig. 4a. Data are represented as means ± SD of two replicates (n = 2).

### Assessing Intra-and inter-assay precision for the TR-FRET assay

Eliminating the wash steps and reducing the overall number of sample handling steps should result in high reproducibility and repeatability. To assess the intra-and inter-assay precision of the TR-FRET assay we selected a set of positive responders as well as negative control samples (68 total) and performed the assay with three operators on three different days (**Fig. 4c, d**). The correlation between operators was above 99.6% with average repeatability of 4.31% and overall precision across days and operators of 5.72%, which well exceeds the commonly desired range for serological assays.

### TR-FRET assay can rapidly be extended to additional antigens

Having established a serological assay for S protein, we set out to assess whether the TRFRET setup is compatible with other antigens. S protein is one of the most widely studied antigens in serological assays for SARS-CoV-2, but there are other SARS-CoV-2 proteins that are highly immunogenic^27^, such as the abundant nucleocapsid protein (N protein) that binds to viral RNA inside the virion^28, 29^. We established an N protein TR-FRET IgG detection assay (thereafter named N TR-FRET). We used the same TR-FRET setup, with the donor fluorophore on the antigen and the acceptor fluorophore on the αIgG antibody. The commercially obtained N protein was biotinylated and terbium-streptavidin conjugate was used to label the antigen. To validate the assay setup, we performed a titration of convalescent CoV2+ serum into biotinylated N protein, Tb-Streptavidin and BODIPY-αIgG. We observed a dose response with strong signal present at lower dilution (1:50), consistent with our S-protein TR-FRET (**Fig. S4**). We next performed N TR-FRET on the 96w_testset which resulted in a sensitivity of 97.6% and specificity 96.6% (**Fig. 5a**). Comparing the S and N TR-FRET readouts on the 96w_testset resulted in a Pearson Correlation coefficient of 0.47, indicating that the two assays are orthogonal and likely provide additive information on serological status when combined (**Fig. 5b**).

**Fig. 5.**
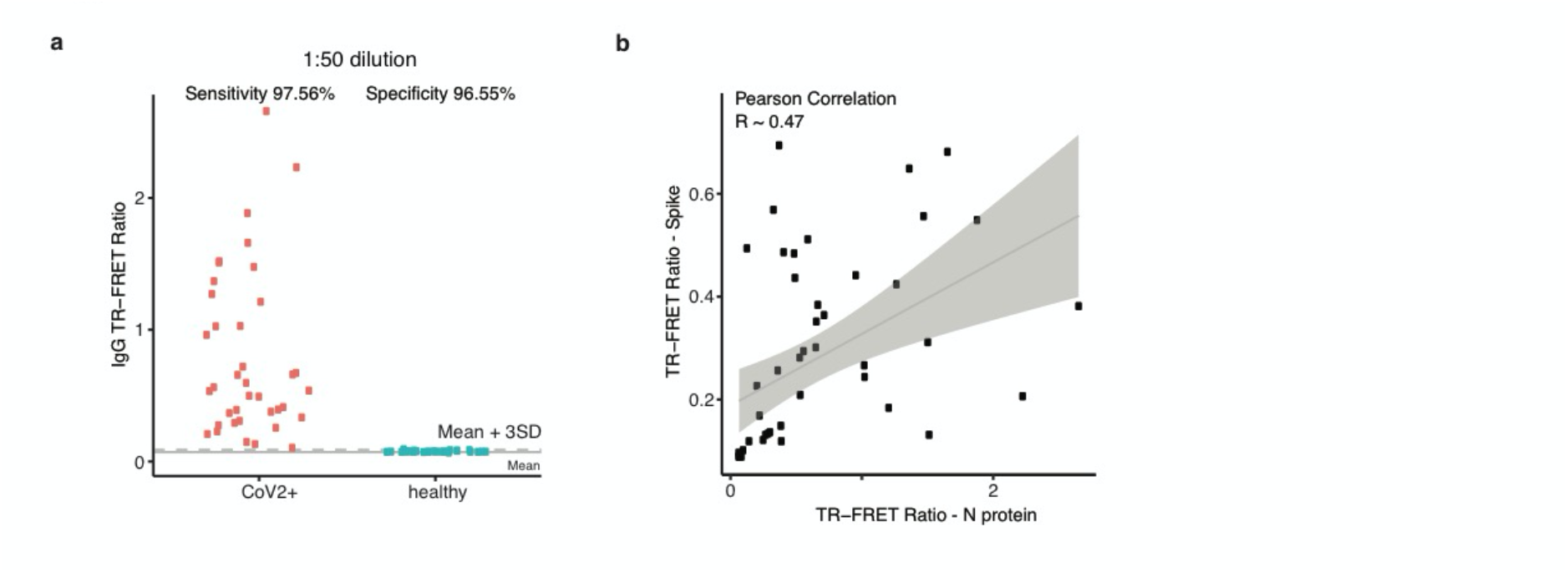
TR-FRET is compatible with other antigens. **a**, Sensitivity and specificity of TR-FRET IgG - N assay performed on a cohort of 45 PCR positive and 30 PCR negative samples (96w_testset). **b**, Correlation of TR-FRET αIgG-S to TR-FRET αIgG-N assays performed on the 96w_testset. Data are represented as means ± SD of two replicates (n = 2).

## Discussion

We describe the development of a homogenous serological assay based on TR-FRET detection, which as we demonstrate provides a scalable alternative to current assay platforms. While assays with signal amplification such as ELISA^30^, or digitized detection such as SIMOA^31^, in theory offer superior detection of low levels of analyte, the TR-FRET assay described here offsets this by low background, allowing for sensitive, accurate detection of SARS-CoV-2 seroconversion. The lack of signal amplification biases the assay towards superior specificity due to very stable background signal, and results in robust reproducibility and repeatability (CV < 5%). When compared to common commercial or ELISA assays on the same samples, the homogenous TR-FRET assay performs equivalent or superior in discriminating SARS-CoV-2^25^.

As we demonstrate with the use of biotinylated N protein, the TR-FRET assay setup offers a flexible platform for rapid onboarding of new antigens of the same pathogen, and can also be easily adapted for testing other pathogens or analytes. The flexibility of ‘in solution’ detection will further allow for future applications such as isotype specific detection, multiple antigens in a single “mix and read” reaction, or the detection of specific epitopes.

In conclusion, the TR-FRET assay fulfills an unmet need for a serological assay that is scalable and has a very low implementation barrier. The scalability arises from the lack of washing steps or sequential manipulations, which lends itself to simple automation using widely available robotic platforms to achieve hundred thousand tests a day. The simplicity of the assay format also makes it easy to implement, as for reproducible results no automated plate washers or similar liquid handling systems are required. With widely available plate readers, one operator could perform several hundred tests a day using manual multichannel pipettes without sacrificing accuracy of the results. Lastly, the miniaturization and lack of large volume wash steps leads to very low costs of the assay.

## Methods

### Antigen production

The full-length Spike protein of SARS-CoV-2 (S protein prefusion stabilized with furin site removed, expressed in TunaCHO) was purchased from LakePharma (Cat. 46328), RBD of SARS-CoV-2 was purchased from LakePharma (Cat. 46438) and full-length biotinylated N protein of SARS-CoV-2 (construct 1-419 with N terminal His-Avi tag) was purchased from Acro biosystem (Cat. NUN-C81Q6).

### Constructs and protein purification

Plasmids encoding CR3022 light chain and IgG, IgM, IgA1 heavy chains were a gift from Galit Alter, MGH, Boston, USA. Antibodies were expressed in Expi293T cells following manufacturer protocol (Thermo Fischer Scientific, A14525) with transfection ratios of 1:1 or 2:1 of heavy to light chain. The cell suspension was cleared using centrifugation, 15 min at 46,500 rcf (Ti45, Beckman Coulter). The clarified media was filtered with 0.45 *μ*m filter before binding to either protein G (GE, GE17-0405-01) for IgG, protein L (GE, GE17-5478-15) for IgM or peptide M (InvivoGen, gel-pdm-5) for IgA1 columns pre-equilibrated with binding buffer (PBS, 10 mM Na_2_HPO_4_, 1.8 mM KH_2_PO_4_, 137 mM NaCl, 2.7 mM KCl at pH 7.4). The beads were washed with 20-50 column volumes (CV) of binding buffer. The protein was eluted from the beads with 6-15 CV of 0.1 M glycine pH 3.0 elution buffer and immediately quenched using a 10:1 ratio of 1 M Tris-HCl pH 8.0. The protein-containing fractions were pooled and flash-frozen in liquid nitrogen at 0.1-1.5 mg/ml. The antibodies were stored at -80°C until further use. Concentrations were estimated using Bradford assay. CR3022 IgG was labeled with BODIPY-NHS as described below.

### Serum samples

Serum/plasma samples used in this study where obtained through the Massachusetts Consortium on Pathogen Readiness (MassCPR) from Mass General Hospital (MGH) and Brigham and Women’s Hospital (BWH), through the Mass General Brigham (MGB) Biobank and through the Ragon Institute. All samples were collected after subjects provided signed informed consent. Four groups of consented subjects were included: 1) hospitalized patients (MGH and BWH) with a SARS-CoV-2 confirmed RNA tests; 2) convalescents patients (MGH) with a confirmed prior SARS-CoV-2 RNA+ and two repeat RNA-negative tests after 2 weeks of isolation; 3) pre-pandemic healthy controls with samples collected prior to December 1, 2019 (MGB Biobank); and 4) a group of low risk community members (Ragon). All samples were tested on an in house validated RBD-specific ELISA as well as on FDA approved Roche and Abbott assays that test for antibodies to N protein. Samples were heat inactivated at 60°C for 1 hour.

### Protein labeling with NCP311-Tb or BODIPY

2.5 ml αhs-IgG (Bethyl, A80-104A), αhs-IgM (Bethyl, A80-100A), αhs-IgA (Bethyl, A80-102A), S protein (LakePharma, 46328) or RBD protein (LakePharma, 46438) at a concentration of 1 mg/ml was buffer exchanged into 100 mM sodium carbonate buffer at pH 8.5, 0.05% TWEEN-20 detergent using PD-10 Desalting Columns (Sigma, GE17-0851-01) according to the manufacturer’s protocol with 0.5 ml per fraction elution. Protein containing fractions were pooled at 0.5 -1 mg/ml and the appropriate volume of either NCP311-Tb (1 mM in dimethylacetamide (DMAc)) or BODIPY-NHS (10 mM in DMSO) was added to achieve a molar ratio of approximately 4-5x NCP311-Tb or 6x BODIPY. The reaction mixture was briefly vortexed and allowed to stand at room temperature for 1 h. To purify the labeled conjugates, the labeling reaction was buffer exchanged into 50 mM sodium phosphate buffer pH 7.4, 137 mM NaCl, 0.05% TWEEN-20 detergent using PD-10 desalting columns following manufacturer protocol using 0.5 ml elution fractions. Protein containing fractions were pooled and flash-frozen in liquid nitrogen at 0.4-0.6 mg/ml concentration and stored at -80°C.

The corrected A_280_ value (A_280,corr_) of protein conjugate was determined via Nanodrop (0.1 cm path length) by measuring A_280_ and A_340_, using equation 1:

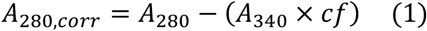

where *cf* is the correction factor for the Tb complex contribution to A_280_ and is equal to 0.157.

The concentration of protein conjugate, *C_ab_* (M) was determined using equation 2:

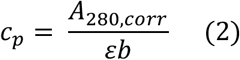

where ε is the antibody extinction coefficient at A_280_, equal to 210,000 M^-1^cm^-1^ for IgG class anti IgG/IgM/IgA Ab, 240,000 M^-1^cm^-1^ for S protein, 80,200 M^-1^cm^-1^ for RBD and *b* is path length in cm (0.1 cm).

The concentration of Tb complex, *C_Tb_* (M) covalently bound to the proteins was determined using equation 3:

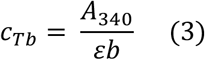

where ε is the complex extinction coefficient at A_340_, equal to 22,000 M^-1^cm^-1^ and *b* is path length in cm (0.1 cm).

The degree of labeling *(DOL)* was calculated using equation 4:

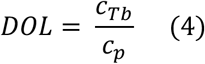

### TR-FRET assay for RBD

Titration of CR3022 IgG/IgM/IgA1 antibody or dilution of tested human serum samples was added to assay mix with final concentrations of 15 nM Tb-labeled RBD, 250 nM BODIPY-labeled αIgG/αIgM/αIgA in a buffer containing PBS, 0.05% Tween-20 (Sigma Aldrich P9416). Serum samples were diluted in the buffer containing 50 mM Tris pH 8.0, 140 mM NaCl, 0.05% Tween-20 and 1% BSA (Cell Signaling Technology 9998S). TR-FRET assays were performed in 384-well microplate (Corning, 4514) with 15 *μ*L final assay volume. Before TR-FRET measurements were conducted, the reactions were incubated for 1h at RT. After excitation of terbium fluorescence at 337 nm, emission at 490 nm (Terbium) and 520 nm (BODIPY) were recorded with a 70 μs delay over 130 μs to reduce background fluorescence and the reaction was followed over >20 or >100 second cycles of each data point using a PHERAstar FS microplate reader (BMG Labtech). The TR-FRET signal of each data point was extracted by calculating the 520/490 nm ratio.

### TR-FRET assay for Spike protein

Titration of CR3022 IgG/IgM/IgA1 antibody or dilution of tested human serum samples was added to assay mix with final concentrations of 7.5 nM Tb-labeled S protein, 250 nM BODIPY-labeled αIgG/αIgM/αIgA in a buffer containing PBS, 0.05% Tween-20 (Sigma Aldrich P9416). Serum samples were diluted in the buffer containing 50 mM Tris pH 8.0, 140 mM NaCl, 0.05% Tween-20 and 1% BSA (Cell Signaling Technology 9998S). TR-FRET assays were performed in 384-well microplate (Corning, 4514) with 15 *μ*L final assay volume. Before TR-FRET measurements were conducted, the reactions were incubated for 1 h at RT. After excitation of terbium fluorescence at 337 nm, emission at 490 nm (Terbium) and 520 nm (BODIPY) were recorded with a 70 μs delay over 130 μs to reduce background fluorescence and the reaction was followed over >20 or >100 second cycles of each data point using a PHERAstar FS microplate reader (BMG Labtech). The TR-FRET signal of each data point was extracted by calculating the 520/490 nm ratio.

### TR-FRET assay for N protein

Dilution of tested human serum samples were added to assay mix with final concentrations of 10 nM biotinylated N protein, 4 nM Streptavidin-Tb and 250 nM BODIPY-labeled αIgG in a buffer containing PBS, 0.05% Tween-20 (Sigma Aldrich P9416). Serum samples were diluted in the buffer containing 50 mM Tris pH 8.0, 140 mM NaCl, 0.05% Tween-20 and 1% BSA (Cell Signaling Technology 9998S). TR-FRET assays were performed in 384-well microplates (Corning, 4514) with 15 *μ*L final assay volume. Biotinylated N protein and Streptavidin-Tb were premixed and incubated for 10 min at RT. Before TR-FRET measurements were conducted, the reactions were incubated for 1 h at RT. After excitation of terbium fluorescence at 337 nm, emission at 490 nm (Terbium) and 520 nm (BODIPY) were recorded with a 70 μs delay over 130 μs to reduce background fluorescence and the reaction was followed over >20 or >100 second cycles of each data point using a PHERAstar FS microplate reader (BMG Labtech). The TR-FRET signal of each data point was extracted by calculating the 520/490 nm ratio.

### ELISA assay for RBD protein

384 well ELISA plates (Thermo Fisher #464718) were coated with 50 *μ*l/well of 500 ng/ml SARS-CoV-2-RBD in coating buffer (1 capsule of carbonate-bicarbonate buffer (Sigma #C3041100CAP) per 100 mL Milli-Q H_2_O) for 30 minutes at room temperature. Plates were then washed 3 times with 100 *μ*l/well of wash buffer (0.05% Tween-20, 400 mM NaCl, 50 mM Tris (pH 8.0) in Milli-Q H_2_O) using a Tecan automated plate washer. Plates were blocked by adding 100*μ*l/well of blocking buffer (1% BSA, 140 mM NaCl, 50 mM Tris (pH 8.0) in Milli-Q H_2_O) for 30 minutes at room temperature. Plates were then washed as described above. 50ul of diluted samples (in dilution buffer; 1% BSA, 0.05% Tween-20, 140 mM NaCl, 50 mM Tris (pH 8.0) in Milli-Q H_2_O) was added to the wells and incubated for 30 minutes at 37°C. Plates were then washed 5 times as described above. 50*μ*l/well of diluted detection antibody solution (HRP-anti human IgG, IgA or IgM; Bethyl Laboratory #A80-104P, A80-100P, A80-102P) was added to the wells and incubated for 30 minutes at room temperature. Plates were then washed 5 times as described above. 40 *μ*l/well of TMB peroxidase substrate (Thermo Fisher #34029) was then added to the wells and incubated at room temperature for 3 minutes (αIgG), 5 minutes (αIgA and αIgM). The reaction was stopped by adding 40 *μ*l/well of stop solution (1 M H_2_SO_4_ in Milli-Q H2O) to each well. OD were read at 450 nm and 570 nm on a Pherastar FSX plate reader.

### ELISA assay for S protein

384-well ELISA plates (ThermoFisher #464718) were coated with 50 μl/well of 500 ng/ml SARS-CoV-2 S protein in coating buffer (1 capsule of carbonate-bicarbonate buffer (Sigma #C3041100CAP) per 100 mL Milli-Q H_2_O) for 30 minutes at room temperature. Plates were then washed 3 times with 100 μl/well of wash buffer (0.05% Tween-20, 400 mM NaCl, 50 mM Tris pH 8.0 in Milli-Q H_2_O) using a Tecan automated plate washer. Plates were blocked by adding 100 μl/well of blocking buffer (1% BSA, 140 mM NaCl, 50 mM Tris pH 8.0 in Milli-Q H_2_O) for 30 minutes at room temperature. Plates were then washed as described above. 50 μl of diluted samples (in dilution buffer; 1% BSA, 0.05% Tween-20, 140 mM NaCl, 50 mM Tris (pH 8.0) in Milli-Q H_2_O) was added to the wells and incubated for 30 minutes at 37°C. Plates were then washed 5 times as described above. 50 μl/well of diluted detection antibody solution (HRP-anti human IgG Bethyl Laboratory #A80-104P) was added to the wells and incubated for 30 minutes at room temperature. Plates were then washed 5 times as described above. 40 μl/well of TMB peroxidase substrate (Thermo Fisher #34029) was then added to the wells and incubated at room temperature for 3 minutes (αIgG). The reaction was stopped by adding 40 μl/well of stop solution (1 M H_2_SO_4_ in Milli-Q H2O) to each well. OD was read at 450 nm and 570 nm on a Pherastar FSX plate reader. The final data used in the analysis was calculated by subtracting 570 nm background from 450 nm signal.

### Statistics

Statistical calculations were performed using Prism 8.0.2 and R v3.6.1; packages ggplot2. The correlation plots include geometrical smoothing using R v3.6.1 geom_smooth function with generalized linear model calculated (glm method) confidence intervals. The samples in ELISA IgG or TR-FRET IgG were classified as positive if the value exceeded the mean (healthy) + 3 SD (healthy) threshold.

## Data Availability

Data is available upon a request.

## Acknowledgments

We would like to thank Dr. Xu Yu and the Massachusetts Coalition for Pandemic Readiness for additional samples provided by the Ragon Institute. This work was supported by a philanthropic gift from *Giving I Grousbeck Fazzalari* (to E.S.F.), by NCI R01CA2144608 (to E.S.F), NSF 1830941 and NIH R21AI13298 (both to R.M.). E.S.F. is a Damon-Runyon Rachleff Investigator (DRR-50-18). N.C.P. is supported by the National Science Foundation Graduate Research Fellowship (DGE1745303). This work was supported by a gift from Ms. Enid Schwartz, by the Mark and Lisa Schwartz Foundation, the Massachusetts Consortium for Pathogen Rediness and the Ragon Institute of MGH, MIT and Harvard.

## Author contributions

E.S.F., H.Y., R.P.N., and D.O. initiated the project, designed experiments, and analyzed data. H. Y., R.P.N., and D.O. performed ELISA assays, developed and performed TR-FRET assays, and expressed and purified proteins and antibodies. N.C.P. and R.M. provided terbium reagents and helped with optimization of assay conditions. S.F., C.A. and G.A. developed ELISA assay, assisted with implementation at DFCI. L.R.B., E.J.N., E.W.K., X.G.Y, J.L., and B.D.W. provided validated serum samples. G.A., R.M., and E.S.F. supervised all aspects of the project and acquired funding. H.Y., R.P.N., and E.S.F. wrote the manuscript with input from all authors. All authors read, revised and approved the manuscript.

